# Distinct temporal stages of infant brain processing associate with early versus later autism diagnosis

**DOI:** 10.1101/2025.01.10.25320053

**Authors:** T. Bazelmans, T. Charman, M. H. Johnson, E.J.H. Jones, BASIS Team

**Author notes:** Correspondence: Tessel Bazelmans, Birkbeck, University of London. Joint senior authors (M. H. Johnson, E.J.H. Jones).

## Abstract

**Background:** The expression of autism traits sufficient to meet criteria for a diagnosis can occur early (by 3 years) or later (from mid-childhood onwards). It remains unknown whether variation in age of onset is due to clinical recognition or whether it reflects distinct biological pathways. One way of addressing this question is by investigating biological differences very early in development associated with age of onset. We use a prospective design to look at event related potentials to faces, one of the most robust biomarkers in autism.

**Methods:** A sample of 102 infants (aged 6-10 months, 54% female) with an older autistic sibling had EEG recorded whilst viewing faces (faces versus noise; gaze towards versus away). Autism diagnostic assessments were conducted at three years and again in mid-childhood (aged 6-12 years), resulting in early diagnosed (at age 3; N=22), later diagnosed (at mid-childhood; N=21) and no autism (N=59) groups.

**Results:** While a short latency response (P1) does not associate with autism outcome, a mid-latency component (N290) associates with early onset autism only, and a later latency component (P400) associates with both early and later onset autism.

**Conclusion:** Temporal stages of face processing in infancy differentially associate with age of autism onset such that an earlier age of diagnosis is associated with earlier stage deviation within the event-related waveform. Early and later onset autism may represent different biological subtypes, with different early brain development, challenging the view of one etiological pathway and that variation in diagnostic age is solely due to clinical ascertainment.

## Introduction

Autism spectrum disorder (hereafter ‘autism’) is a neurodevelopmental condition that affects 1-2% of children (Maenner et al., 2023; Roman-Urrestarazu et al., 2021). There is extensive heterogeneity across the autism spectrum in core autistic traits and in the presence and impact of common co-occurring conditions. Variation is present both between individuals with a diagnosis and within individuals across the lifespan (Lord et al., 2022). This diversity may reflect underlying causal genetic and / or brain differences (Jeste & Geschwind, 2014), as well as risk and protective effects of differential environmental exposures (Molnar-Szakacs et al., 2021). Consequently, there is growing interest in stratification of autism into sub-types that may map better on to underlying biomarkers, with resulting implications for improving early identification and support (Loth et al., 2016).

One of the ways in which autism could be stratified is by age of onset and this might explain some of the heterogeneity in core and co-occurring trait expression (Zhang et al., 2024). This aligns with the ‘chronogeneity’ concept introduced by Georgiades et al. (2017) to describe the developmental time course of individual trajectories of emergence. Although a reliable diagnosis can be made in some children from 2 years (Ozonoff et al., 2015), the age at which autism symptoms manifest can vary from toddlerhood to mid-childhood and beyond; traits of autism may also not be clearly recognised until social demands exceed limited capacities (DSM-5; American Psychiatric Association, 2013). In addition to variation in the onset of symptoms, there is usually a delay between parents and teachers reporting difficulties and receiving a community diagnosis of autism (Hosozawa et al., 2020). Many children do not receive a diagnosis until they reach school age (Brett et al., 2016; Hosozawa et al., 2020) and some may not receive a community diagnosis until adulthood (Russell et al., 2021). Contextual factors such as access to services, variation in local practice and under-recognition of autism particularly when other conditions are present (‘diagnostic overshadowing’; Davidovitch et al. (2023)) impact age of community recognition and diagnosis (Hosozawa et al., 2020; O’Nions et al., 2023). These systematic factors that influence the timing of receiving a clinical diagnosis in the community make it difficult to assess the age of symptom onset in general population and clinically recruited autism samples. Thus, to identify whether the temporal manifestation of autism is truly a stratification factor requires prospective longitudinal studies in which autism traits are assessed at multiple age points from the early years.

Prospective longitudinal studies of ‘infant sibs’ (babies with a family history of autism, most commonly an older sibling with a diagnosis) have shown that around 20% of infant sibs meet criteria for autism by age 3 years, consistent with the highly heritable nature of autism (Ozonoff et al., 2024). However, few such studies have followed these infants through to mid-childhood and later (Miller et al., 2016; Shephard et al., 2017). A recent study followed a cohort of infant sibs to mid-childhood with autism diagnostic assessments conducted at 3 years and again in mid-childhood. Strikingly, twice as many children at increased familial likelihood for autism were diagnosed with autism at mid-childhood compared to 3 years (Bazelmans et al., 2024). Of those later diagnosed, approximately half showed no symptoms and had no reported parental concerns at the earlier assessment. Thus, there is a considerable proportion of infants with a family history of autism who do not manifest clinically relevant autism traits until later in development.

Infant sib studies have also shown that a range of facets of infant brain development differ in children who later receive an autism at 2-3 years (Dawson et al., 2023; Szatmari et al., 2016). For example, several studies have shown that electrophysiological differences in response to faces are already present in infants with a family history of autism who meet autism criteria as toddlers (Elsabbagh et al., 2012; Jones et al., 2016; Tye et al., 2022). Event-related potentials (ERP) consist of a series of deflections representing coordinated neural activity that reliably occurs time-locked to stimulus presentation, reflect different stages of information processing, and develop in order from lower to higher processing levels over early development (Whitehead et al., 2019). Early event related potentials such as the P1 (a positive-going deflection over the visual areas that occurs after around 100ms) primarily reflect sensory processing. Mid-latency components, such as the N170 in adults or N290 in infants (negative going deflections over temporal channels at the indicated latencies) reflect more structural decoding and show greater sensitivity to faces than objects or other nonsocial stimuli (De Haan et al., 2003). Longer latency components, such as the P400, reflect later stage processing (De Haan et al., 2003). The most replicated electrophysiological marker of autism is a slower N170 response to faces (Kang et al., 2018), which has become the focus of biomarker studies such as the Autism Biomarkers Consortium for Clinical Trials (ABC-CT) (McPartland et al., 2020) and LEAP (Mason et al., 2022). In clinical samples, these atypicalities in face processing have been observed from the age of three (Dawson et al., 2005, 2023). Within prospective studies from infancy, infants without later autism show longer N290 latencies to faces versus noise, whereas the autism group showed no differentiation (Tye et al., 2022). Shorter latency to faces versus noise was also associated with reduced socialisation skills (Tye et al., 2022) and a higher autism polygenic score (Gui et al., 2021). Further, Jones et al. (2016) showed that 6-month-old infants with a diagnosis at 3 years showed faster P400 responses to faces compared to those without a diagnosis; shorter P400 latency was associated with more autism symptoms at 24 months. We have also reported that in response to dynamic gaze shifts, 8-month-old infants diagnosed with autism at age 3 years showed longer P1 and P400 latencies to gaze shifting towards versus away in infancy, whereas the no-autism and typical likelihood children showed the opposite pattern (Elsabbagh et al. 2012; Tye et al., 2022). Thus, neural responses to faces in infancy are altered in children with an early diagnosis of autism.

In the current study we test whether infant neural responses to faces are sensitive to diagnostic timing in a prospective cohort, which would be consistent with the proposal that different timing of symptom manifestation represents different biological trajectories. Specifically, we examine infant face processing ERP responses in those children in our extended prospective family history study (Bazelmans et al., 2024) who did not receive a diagnosis until mid-childhood (later-autism). We compare this later-autism group to children who did not receive an autism diagnosis at either visit (no-autism) to determine whether there are any detectable infant neural face processing differences in children with a later diagnosis; and to those diagnosed at age 3 years (early-autism) to assess whether there are differences in infant neural face processing related to diagnostic timing. Of note, we have already examined the relation of infant ERP responses to 3 year diagnosis in this cohort (Gui et al., 2021; Tye et al., 2022). Specifically, based on previous work we preregistered hypotheses regarding modulation of the latency of the N290 to faces vs. noise (Gui et al., 2021; Tye et al., 2022) and modulation of the latency of the P1 and P400 to gaze shifting towards vs. away from the infant (Tye et al., 2022). If early vs. later-diagnosed cases represent distinct subgroups, they should significantly differ from each other and from children without a later diagnosis on these components. Lastly, we hypothesise that any infant ERP features that associate with later diagnosis (eg shorter N290 to face vs. noise and longer P1 and P400 to gaze towards vs. away) should be associated with higher social communication and restricted and repetitive autistic traits in mid-childhood given evidence of shared aetiology between dimensional traits and categorical diagnosis (Robinson et al., 2016; Ruzzo et al., 2019) (for additional details on our hypotheses, also see pre-registration (link below) and Supplementary Materials).

## Methods & Materials

The hypotheses and analysis plan for this manuscript are pre-registered on Open Science Framework (OSF; https://doi.org/10.17605/OSF.IO/XSYU7). We have noted in text where we have deviated from the pre-registered analysis plan.

### Participants

The data is part of Phase 1 and Phase 2 of the longitudinal British Autism Study of Infant Siblings (BASIS; www.basisnetwork.org) (see Figure 1 for study design), which follows infants with a familial likelihood of autism (because of an older sibling with a diagnosis of autism) and those with an older sibling but without a first-degree family member with autism (typical likelihood) through multiple timepoints in early development (8, 14, 24 and 36 months). In total, 159 elevated likelihood infants were followed up in mid-childhood (6 years 9 months – 12 years 8 months) (Bazelmans et al., 2024). We excluded three infants because they received a research diagnosis at 3 years, but not at mid-childhood (Shephard et al., 2017). Of the remaining children, 102 had EEG data at infancy and a diagnostic outcome at mid-childhood available: 59 (22 male) No-autism, 22 (15 male) Early-autism, and 21 (10 male) Later-autism (Gender: χ^2^(2) = 6.18, p = .045). Three of the children only had a half-sibling with a diagnosis. To note, we have repeated all analysis excluding half-siblings in line with the pre-registration, but results remained the same.

**Figure 1.**
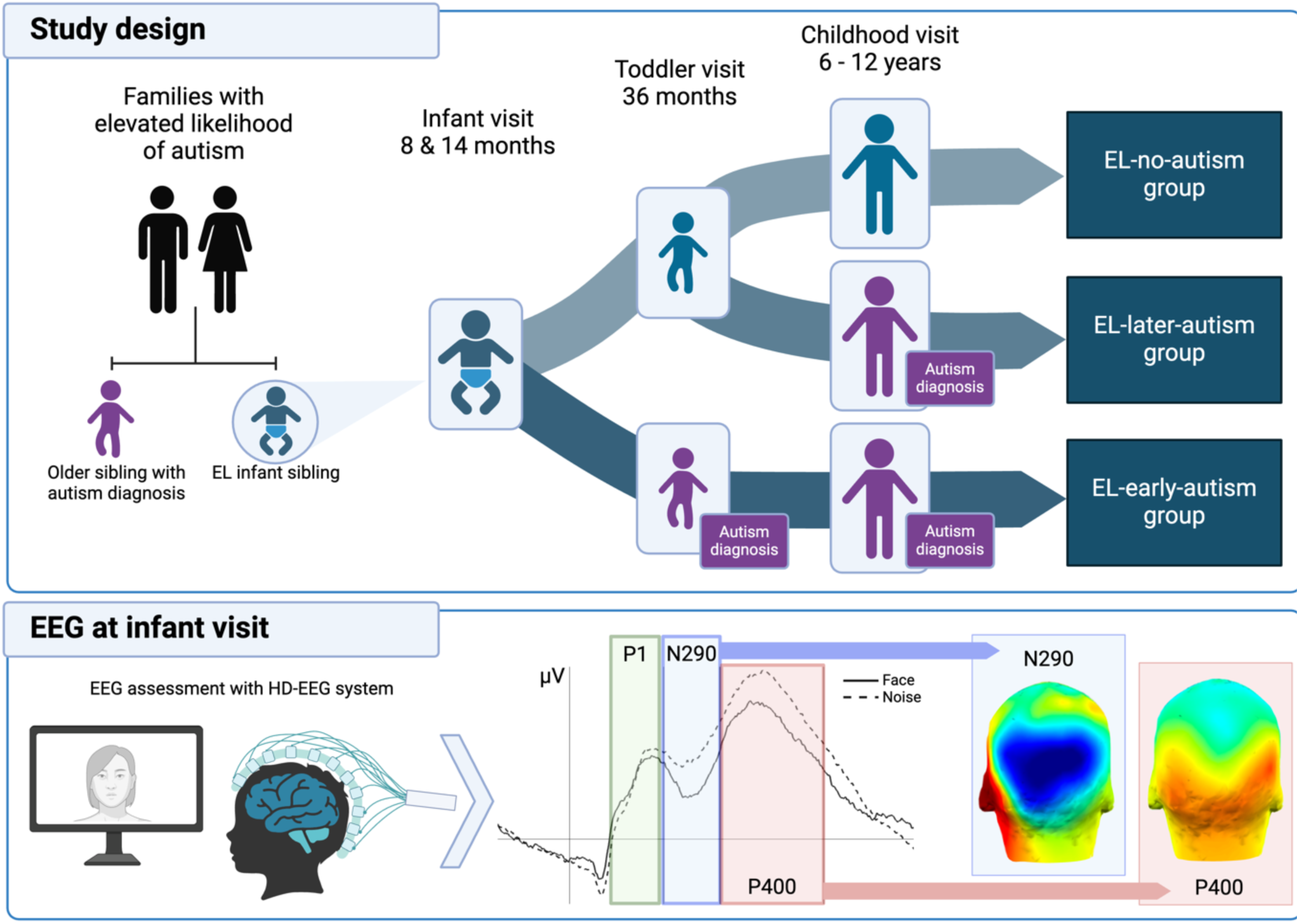
Study design. For stimuli used in the EEG task see Elsabbagh et al. (2012).

### Electrophysiological Measures (8 months)

The task was as reported by Elsabbagh et al. (2012) and Tye et al. (2022)(also see Supplemental Materials) and was designed to assess electrophysiological responses to: 1) faces (static and irrespective of gaze direction) versus visual noise (N290) and 2) dynamic gaze shifts (gaze towards versus away from the infant; P100 & P400). EEG was recorded from a 128 channel Hydrocel Sensor Net. Following artifact detection and rejection (see online supplemental materials), stimulus-locked epochs (−200 to 800ms peristimulus window) were averaged for each of the three contrasts. Peak amplitude and latency of the averaged P100, N290, and P400 across occipitotemporal channels for each stimulus/contrast were used as input features for subsequent analyses (Elsabbagh et al., 2012; Tye et al., 2022).

### Clinical Assessments (age 3 years and mid-childhood)

Experienced researchers and clinicians (including TB, TC) determined the best estimate clinical outcome at both 3 years and mid-childhood by reviewing all available information from visits using ICD-10 (Phase 1: 3 years) or DSM–5 (Phase 1: mid-childhood and Phase 2) criteria. This information included (but was not limited to) the Autism Diagnostic Interview – Revised (ADI-R)(Lord et al., 1994), Autism Diagnostic Observation Schedule (ADOS; Lord et al., 2012), Vineland Adaptive Behavior Scale 2^nd^ (VABS-II) (Sparrow et al., 2005) or 3^rd^ edition (VABS-3) (Sparrow et al., 2016), Mullen Scales of Early Learning (Mullen, 1995) (24 & 36 months) and Wechsler Abbreviated Scale of Intelligence (Wechsler, 2011)(mid-childhood). Researchers were not blind to either likelihood group or to 36 months best estimate clinical outcome (see Bazelmans et al., 2024 for details).

### Social Communication and Restricted Interest and Repetitive Behaviours (mid-childhood)

Parents reported on their child’s autism traits by completing the Social Responsiveness Scale (SRS-2)(Constantino & Gruber, 2012). This questionnaire consists of 65 items, rated on a four-point Likert scale, ranging from not true to almost always true. Two main subscales can be calculated: the Social Communication and Interaction (SCI; 53 items) and the Restricted Interests and Repetitive Behaviour (RRB; 12 items) subscales.

### Statistical Analysis

All analysis were performed in Stata 18 (StataCorp, 2023). Mixed models were run (*`mixed`* command, followed by *`contrast`* command), including the random intercept for participant ID, using restricted maximum likelihood (REML). In the first model, diagnostic outcome (No-autism, Early-autism, Later-autism), condition (face vs. noise / towards vs. away) and their interaction effect were included. The models were repeated including Phase, gender, age and non-verbal developmental score (NVT) at the 8-month visit. Model fit of the mixed models and the ANOVAs was checked by inspecting the normality of standardised residuals and models were repeated excluding any standardised residuals >|3|. Any significant interaction was further explored by running a post-hoc ANOVA on the difference score (i.e. the difference between the two conditions). Our hypotheses rest on the contrasts between the Late-autism and Early-autism/No-autism groups and are corrected for these two comparisons. The contrast Early-autism versus No-autism is reported for completeness, but were previously reported (Tye et al., 2022).

To examine bivariate associations between infant ERP (P1, P400 and N290 difference scores) and mid-childhood autism traits (SRS-SCI and SRS-RRB), we ran separate Kendall Tau_b_ correlations. We planned to conduct multivariate regression. However, due to the skewness of the SRS data these models did not lead to a good fit, even after transformation, and so we used non-parametric tests instead and corrected for multiple comparisons (n=6). Significant correlations were repeated as ordinal regressions with SRS scores as outcome variable including also Phase, gender, and age and NVT at the infant visit. Dimensional associations with the ADI-R and ADOS-2 scores are reported in the Supplementary Materials.

## Results

Descriptive statistics can be found in Table 1. As previously reported in Bazelmans et al. (2024), the Later-autism group showed lower Vineland and higher SRS scores at mid-childhood compared to the No-autism group, but similar to the Early-autism group (indicating no clear differences in severity based on early vs late diagnosis). ERP descriptives can be found in Table 2 and are visualised in Figure 2.

**Figure 2.**
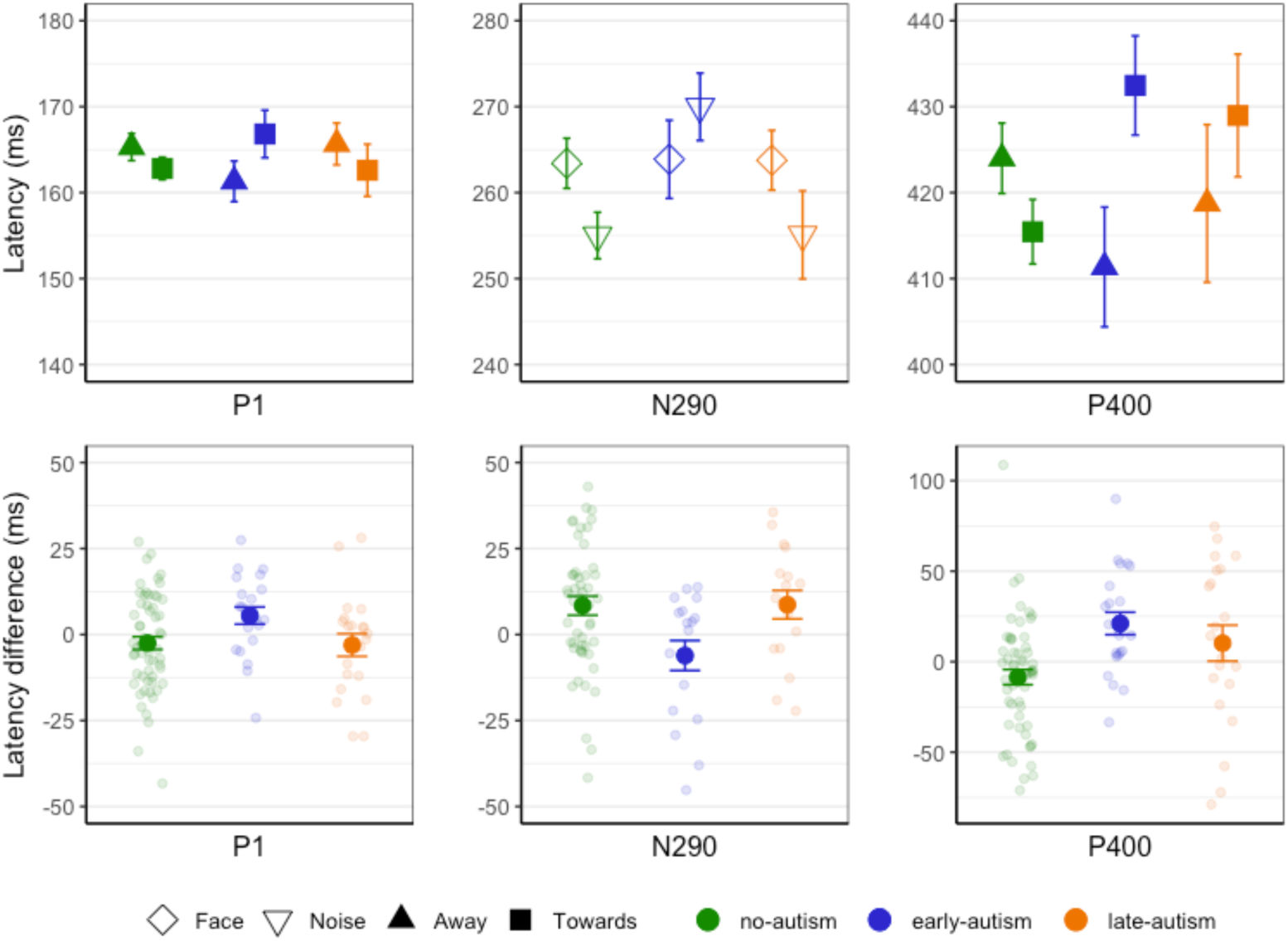
Mean and SE of latency responses by ERP and mid-childhood outcome group.

**Table 1:**
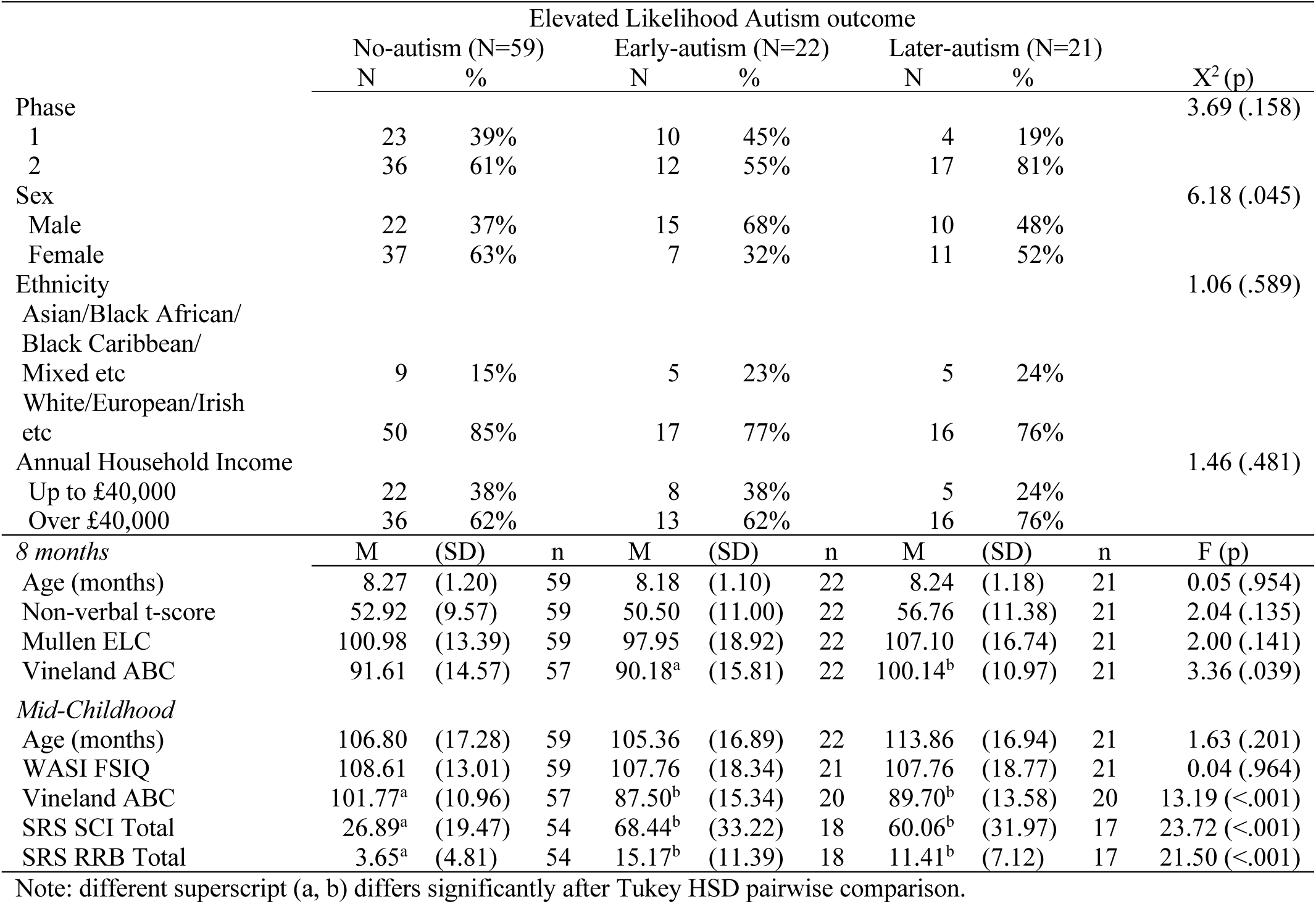
Descriptive statistics by outcome group.

**Table 2:** 8-month-old infant ERP latencies by mid-childhood outcome group.

|  | No-autism (N=59) |  |  | Early-autism (N=22) |  |  | Later-autism (N=21) |  |  |
| --- | --- | --- | --- | --- | --- | --- | --- | --- | --- |
|  | M | (SD) | n | M | (SD) | n | M | (SD) | n |
| *P1 Latency* |  |  |  |  |  |  |  |  |  |
| Shift Towards | 162.81 | (10.11) | 59 | 166.82 | (12.97) | 22 | 162.60 | (13.89) | 21 |
| Shift Away | 165.30 | (12.12) | 59 | 161.31 | (11.08) | 22 | 165.66 | (11.16) | 21 |
| Towards-Away | -2.49 | (14.17) | 59 | 5.51 | (11.71) | 22 | -3.06 | (15.07) | 21 |
| *N290 Latency* |  |  |  |  |  |  |  |  |  |
| To Faces | 263.41 | (20.81) | 51 | 263.87 | (19.28) | 18 | 263.77 | (14.33) | 17 |
| To Noise | 255.01 | (19.33) | 51 | 269.98 | (16.64) | 18 | 255.09 | (21.12) | 17 |
| Faces-Noise | 8.40 | (19.81) | 51 | -6.11 | (18.43) | 18 | 8.68 | (17.03) | 17 |
| *P400 Latency* |  |  |  |  |  |  |  |  |  |
| Shift Towards | 415.45 | (28.57) | 58 | 432.46 | (27.08) | 22 | 428.97 | (32.69) | 21 |
| Shift Away | 423.99 | (31.18) | 58 | 411.35 | (32.67) | 22 | 418.74 | (42.03) | 21 |
| Towards-Away | -8.54 | (31.80) | 58 | 21.11 | (29.10) | 22 | 10.23 | (45.52) | 21 |

*P1 Latency*: The overall model was not significant, indicating that group and condition did not explain significant variance (Wald χ^2^(5) = 6.38, p = .271).

*N290 Latency*: The overall model was significant (Wald χ^2^(5) = 18.31, p = .003). There was no main effect of group (p = .227) or condition (p=.115) but there was a significant group by condition interaction effect (χ^2^(2) = 8.36, p = .015). The posthoc ANOVA was significant (F(2,83) = 4.18, p=.019, *η*^2^ = .09) and pairwise comparison showed that the Later-autism group had longer latencies to face versus noise compared to the Early-autism group (p = .024, after correction for two comparisons: p = .048), who showed longer latencies to noise versus face. The Later-autism and No-autism groups did not differ (p = .958). As previously reported (Tye et al., 2022), the No-autism and Early-autism groups significantly differed from each other (p = .007). None of the control variables were significant predictors (all p > .094).

*P400 Latency*: The overall model was significant (Wald χ^2^(5) = 14.02, p = .015). There was no main effect of group (p = .818) but there was a significant effect of condition (χ^2^(1) = 3.95, p = .047) and a group by condition interaction (χ^2^(2) = 13.34, p = .001). The posthoc ANOVA was significant (F(2,98) = 6.67, p = .002, *η*^2^ = .12). Pairwise comparison showed that the Later-autism group had longer P400 latencies to faces shifting towards versus away, compared to the No-autism group (p = .035, after correction: p = 0.07), who showed the opposite pattern. The Later-autism group did not differ from the Early-autism group (p = .304). As previously reported (Tye et al., 2022), the Early-autism also showed longer P400 latencies to faces towards versus away compared to the No-autism group (p = .001). Results became more pronounced after removing one outlier in the ANOVA (later-autism versus no-autism: p = .014, after correction: p = .028). None of the control variables were significant predictors (all p > .185).

### Associations with autism traits

There was a significant association between P400 latency difference score in infancy and both social-communication impairment (Ktau = .200, *p* = .005, corrected: p = .03) and restricted and repetitive behaviour (Ktau = .214, *p* = .003; corrected: p = .018) SRS subscales in mid-childhood. Specifically, slower responses to gaze turning towards vs. away were associated with higher autistic traits in mid-childhood. These associations remained significant (SCI p = .003 and RRB p = .004) after including the control variables (all *p* > .225). No associations were found between SCI or RRB traits in mid-childhood and either P1 (*p* = .324 and p = .984, respectively) or N290 (*p* = .156 and p = .504) latency difference scores.

## Discussion

This is the first study to identify differential infant brain processing markers for children who received an early versus later diagnosis of autism, and the first to show that there are differences in the infant brain in children who are diagnosed with autism after toddlerhood. We compared 8-month-old infant ERP responses to face stimuli of infant sibs with an autism diagnosis in toddlerhood versus in mid-childhood, and infants without later autism. We found that different stages of brain processing of faces in infants differentially associate with age of symptom manifestation. Specifically, while a short latency component (P1) did not differentiate between elevated likelihood groups, mid-latency face processing (N290) was different only in children with early diagnosis, and a longer latency face processing component (P400) was different in children with both early and later diagnosis relative to infants without an autism diagnosis.

We tested whether the Later-autism group differed on ERP latencies from the No-autism group, reflecting that underlying brain differences would already be evident in infancy even if phenotypic expressions did not (yet) meet diagnostic criteria by the age of 3 years. While children with a later autism diagnosis showed differences in P400 latency, they did not for the earlier N290 component. Further, the Later-autism group responded similarly to the Early-autism group on the P400 latency component, showing that the early- and later-autism groups have similar neurodevelopmental differences in later-stage face processing. However, for the earlier N290 latency response the Later-autism group were significantly different from the Early-autism group, instead resembling children who did not meet criteria for autism at either timepoint (No-autism). We also found dimensional associations between the latency of the infant P400 response and mid-childhood autism trait severity both for social communication difficulties and for restricted, repetitive and rigid behaviours, suggesting that later, mid-childhood trait measures of autism severity, as well as categorical autism diagnosis, are associated with differences in later stage electrophysiological components.

The findings imply a parallel between the stage of real-time brain processing of faces and the timing of the developmental manifestation of autism traits. The earlier behavioural symptoms become evident, the earlier in the ERP waveform atypicalities in information processing occur, suggesting disruptions in relatively early-stage information processing. Differences in those with and without an autism diagnosis at age 3 years became evident from early stages of face-specific structural/configural processing onwards. The N290, mostly recorded by posterior electrodes over the temporal lobe area like the adult N170, is thought to reflect the encoding of structural information in faces (De Haan et al., 2003; Halit et al., 2003). We have previously shown that infants with Early-autism show faster N290 responses to face versus noise at 8 months (Gui et al., 2021; Jones et al., 2016; Tye et al., 2022). Further, faster and larger responses to neutral faces in three-year-olds have also previously been associated with lower ADOS symptom severity scores in adolescents (Neuhaus et al., 2016). The N290 is considered a precursor of the adult N170 response, which has repeatedly been found to be delayed in autism (Kang et al., 2018). In a subsample of our current sample, we previously reported larger N170 amplitude, but not latency, in elevated likelihood children in mid-childhood compared to typical likelihood children (Shephard et al., 2020). Given we show that the infant N290 only varies in children with an early diagnosis and as the N170 has been considered as a biomarker for autism (Mason et al., 2022; McPartland et al., 2020), it will be important to evaluate whether considering chronogeneity in the form of age of onset would improve the use of the N170 as a stratification biomarker.

Children who received a diagnosis of autism at mid-childhood, but not at age 3 years, show mainly disruptions in the P400. The P400 occurs around 400ms after stimulus onset and reflects mid to higher-level, semantic, information processing, and is mostly observed in posterior and lateral regions (De Haan et al., 2003). Like the Early-autism group, the Later-autism group showed longer P400 latencies to faces shifting gaze towards versus away, but this was opposite from the No-autism group (a finding similar to our exploratory work in Bedford et al. (2017)). We did not find any significant group effects of the P1 latency. This despite our previous report of P1 difference in infants later diagnosed with autism at 3 years (Tye et al., 2022), but in line with other reports (McCleery et al., 2009). Early ERP responses, such as the P1 are associated with early sensory processing, peaking around the occipital area, and responses are usually not specific to faces in infants. Thus, whereas basic sensory processing seems relatively typical, issues with more structural processing (N290) are present in those with early autism, and children diagnosed later might have particular issues with integrating the sensory and perceptual input reflected in P400 changes (Li et al., 2024).

No group effects of the amplitude models were found (see Supplementary Materials). Although various studies have suggested differences in amplitude in infants later diagnosed with autism (Elsabbagh et al., 2012), these effects have not consistently been reported to be specific to later diagnosis and instead are found to relate to autism family history status (Tye et al., 2022). Also, larger N290 amplitude to face versus noise in infants related to more social-communication problems in mid-childhood, however his effect was driven by atypical processing of noise rather than faces (Shephard et al., 2020).

Together our evidence supports an intriguing association with increasing effects as real-time neural processing takes place. One possibility is that subtle differences in excitation/inhibition balance may lead to signal-to-noise changes whose effects become increasingly compounded as a sequence of neural processing proceeds (Faisal et al., 2008; Hancock et al., 2017). Smaller deviations from neurotypical brain processing result in ERP differences that appear later in the waveform, and a later age of diagnosis. Another possibility is that that there are distinct biological pathways underlying an early versus later autism diagnosis. This idea follows recent findings of Zhang et al. (2024), showing age of autism diagnosis is associated with different (though partially correlated) sets of common genetic variants.

One of the limitations to note is that our data did not allow us to look more closely at the specific age symptoms manifested but rather those meeting criteria at 3 years versus 6-to-12 years, the two timepoints at which we conducted autism diagnostic assessments. This limited our ability to look at dimensional association between age and where differences in the EEG waveform emerge. Moreover, our hypotheses and analyses were driven by earlier findings within cohorts overlapping with the current sample; expanding analysis to other features that were not previously examined in relation to early autism may identify new brain changes associated with solely later diagnosis, and replication in independent samples is necessary. Nevertheless, our prospective study provides a unique design where we can combine infant EEG with longitudinally reported symptoms in a cohort in which diagnosis is not influenced by demographic or societal factors that affect the age at which community diagnoses are received from clinical services (Hosozawa et al., 2020; Russell et al., 2021).

To conclude, our findings on the association between stage of real-time brain processing and age of symptom onset, together with recent genetics evidence for variation in age of diagnosis (Zhang et al., 2024), support the view that the autism spectrum could be stratified by ‘chronogeneity’ – informative variance about the developmental time course encompassing individual trajectories of emergence (Georgiades et al., 2017). Understanding mechanistic and biological differences related to age of onset could bring new insights and provide better targeted developmentally sensitive intervention and support for autistic individuals. Further research should explore age of emergence as a stratification marker.

## Data Availability

All data produced in the present study are available upon reasonable request to the authors

## Acknowledgements

The authors are grateful to all the families who took part in this study. The BASIS Team consists of: Jannath Begum Ali, Rowan Arthur, Jeni Baykoca, Anna Blasi, Patrick Bolton, Celeste Cheung, Kenny Chiu, Leila Dafner, Kim Davies, Mayada Elsabbagh, Janice Fernandes, Laurel Fish, Isobel Gammer, Teodora Gliga, Jeanne Guiraud, Rianne Haartsen, Sarah Kalwarowsky, Anna Kolesnik, Michelle Liew, Sarah Lloyd-Fox, Helen Maris, Luke Mason, Mia Medas, Bosiljka Milosavljevic, Louise O’Hara, Greg Pasco, Andrew Pickles, Laura Pirazzoli, Helena Ribeiro, Erica Salomone, Elizabeth Shephard, Chloë Taylor, Leslie Tucker, Charlotte Tye. For the purposes of open access, the author has applied a Creative Commons Attribution (CC BY) license to any Accepted Author Manuscript version arising from this submission. We are grateful to Rianne Haartsen for producing Figure 1 (Created in BioRender. Haartsen, R. (2025) https://BioRender.com/h54c434) and to Charlotte Tye for assistance with processing and analysis of EEG data.

## Ethical Statement

Ethical approval was obtained from the NHS National Research Ethics Service (NHS RES London REC 06/MRE02/73, 08/H0718/76 and 14/LO/0170) and PNM Research Ethics Subcommittee, King’s College London (RESCM-18/19-10556). Parents gave informed consent to participate in the study.

## Author Contributions

Conceptualization: Bazelmans, Charman, Johnson, Jones

Investigation: Bazelmans

Data Curation: Bazelmans

Formal Analysis: Bazelmans

Funding Acquisition: Johnson, Charman, Jones

Writing – Original Draft: Bazelmans

Writing – Review and Editing: Bazelmans, Charman, Johnson, Jones

