## Supplemental Materials for "Distinct temporal stages of infant brain processing associate with early versus later autism diagnosis"

### Stages of brain processing in infants associate with early versus later autism diagnosis – Supplementary Materials

### Pre-registered hypotheses

Our detailed hypotheses, including hypotheses related to visual disengagement and alpha-connectivity (not reported in this manuscript) were pre-registered on Open Science Framework (OSF; https://doi.org/10.17605/OSF.IO/XSYU7). Here we report the hypotheses related to the EEG/ERP data only:

We hypothesise that the late diagnosis group has a similar early-emerging neurodevelopmental difference that underlies their eventual diagnosis of autism by mid-childhood as the early diagnosis group but that this might differ by degree or severity such that for the late diagnosis group the emerging behavioural atypicalities are insufficient for them to meet diagnostic criteria at the age of 3 years (e.g., subthreshold). In line with our hypothesis that the late diagnosis group has underlying (but less severe) early-emerging neurodevelopmental differences, we expect the EL-late-autism group to be significantly different from the EL-no-autism group (H1). Further, we test two possible predictions:

• the EL-late-autism group does not differ from the EL-early-autism group on neural/cognitive measures in infancy. That is, the early and late groups are similar at a neural/cognitive level, despite the EL-late-autism group not meeting diagnostic criteria on a behavioural level at 3 years and only doing so in mid-childhood (H2.1); or

• the EL-early-autism group has a more severe manifestation of autism terms of neural/cognitive differences compared to the EL-late-autism group. This underlies their higher behavioural atypicality that reaches the threshold for diagnosis at 3 years and their higher severity of autism symptoms in mid-childhood.

Based on this theory, we expect a graduation of difference in infancy such that the EL-late autism group will significantly differ from the EL-early-autism group. The direction would be such that the EL-late-autism group has measures closer to those without autism and the EL-early-autism group being most ‘atypical’ (H2.2).

For the ERPs specifically, we hypothesize that:

- N290 latency: (i) the EL-late-autism group will show shorter latencies to face versus noise compared to the EL-no-autism group (H1) (ii) and the EL-late-autism group will have latencies similar to the EL-early-autism group, i.e. no differentiation between noise vs face on N290 latency (H2.1) or will show longer latencies to face versus noise compared to the EL-early-autism group (H2.2)(but not as long as the EL-no-autism group).
- P1 + P400 latency: (i). the EL-late-autism group will show P1/P400 latencies that are significantly different from the EL-no-autism group, who will show longer latencies to gaze shift away versus towards (H1). (ii) and the P1/P400 latencies of the EL-late-autism group will be either similar to the EL-early-autism group, i.e. latencies are longer to gaze

shift towards versus away (H2.1) or the contrast between towards versus away will be smaller compared to the EL-early-autism group (H2.2)(but still different from the EL-no-autism group).

- Hypothesis 3: Based on the observed differences in infant measure in association with outcome groups (see Hypothesis 1 & 2) and previous findings of associations with later traits, we expect the infant measures show an association with autism traits at mid-childhood. Specifically, we expect: ERP measures to be associate with traits in the social communication domain, especially faster N290 response to noise versus face and faster P1 response to shift away versus towards associated with better social skills (Tye et al., 2022) and larger N290 response to face versus noise to be associated with higher social communication problems (Shephard et al., 2020).

### Additional EEG methods

Methods are as reported in Elsabbagh et al. (2012) and Tye et al. (2022). Infants sat on their parents' laps at a 60 cm distance from a 40 x 29 cm computer screen. Gaze during stimulus presentation was recorded by video camera. Each trial block began with a static colorful fixation stimulus followed by a color image of one of four female faces, with gaze directed either toward or away from the infant. In subsequent trials of the same block, the face remained on the screen but displayed three to six gaze shifts, alternating from directed toward to away from the infant. Faces were aligned with the center of the screen with the eyes appearing at the same location as the fixation stimuli, to ensure that infants were fixating the eye region. The faces subtended 21 x 14 degrees of visual angle. In addition to face trial blocks, during approximately one third of all blocks, infants were presented with ''visual noise'' stimuli. The latter were constructed from the same faces presented within the task, by randomizing the phase spectra while keeping the amplitude and color spectra constant. Fixation stimuli, preceding the onset of the face and noise stimuli, subtended approximately 1.6 3 1.6 degrees and were presented for a variable duration of 800 to 1,200 ms. Each trial lasted for 1,000 ms. A 128 channel Hydrocel Sensor Net was mounted on each infant's head, while they were seated on the parent's lap in front of the stimulus screen. When the infant was attending toward the screen, trials were presented continuously for as long as the infant remained attentive, with brain electrical activity measured simultaneously using the vertex as a reference (Cz in the conventional 10/20 system). EGI NetAmps 200 was used (gain = 1,000). Data were digitized with a sampling rate of 500 Hz and band-pass filtered between 0.1–100 Hz.

Data were stored and analysed offline in EGI Netstation version 5.2.0.2 (using the same protocol as Elsabbagh et al. (2012))). Trials were retained only when infants were fixating on the centre of the screen at stimulus onset, without any gaze shifts, blinking or head movements during the 800ms segment following stimulus onset. Data were then corrected to the -200ms baseline. Following automated artifact detection, an experienced EEG researcher (CT). conducted detailed manual artifact rejection through visual inspection of individual trials. Data from any sensor were excluded if they contained artifacts. Missing data from 12 or fewer channels were interpolated, otherwise the entire trial was rejected. Data were then rereferenced to the average.

Stimulus-locked epochs (-200 to 800ms peristimulus window) were averaged for the following trial contrasts: (1) faces (valid static (irrespective of gaze direction) vs. visual noise stimuli presented at the beginning of each block); and (2) dynamic gaze shifts (gaze toward vs. away from the infant, after appearance of the initial face within each block). Averages were computed for each participant in each condition on a minimum of 10 trials. Due to variable rates of presentation of each stimulus type, a different number of trials were included for each contrast, which did not differ by outcome group. The occipito-temporal montages from Elsabbagh et al. (2012)were used (Figure S1) and corroborated with visual inspection of the grand average for each condition across the three contrasts. Peak amplitude and latency of the average P1, N290 and P400 responses were included in subsequent analyses because consistently modulated in face processing tasks in the first year of life.

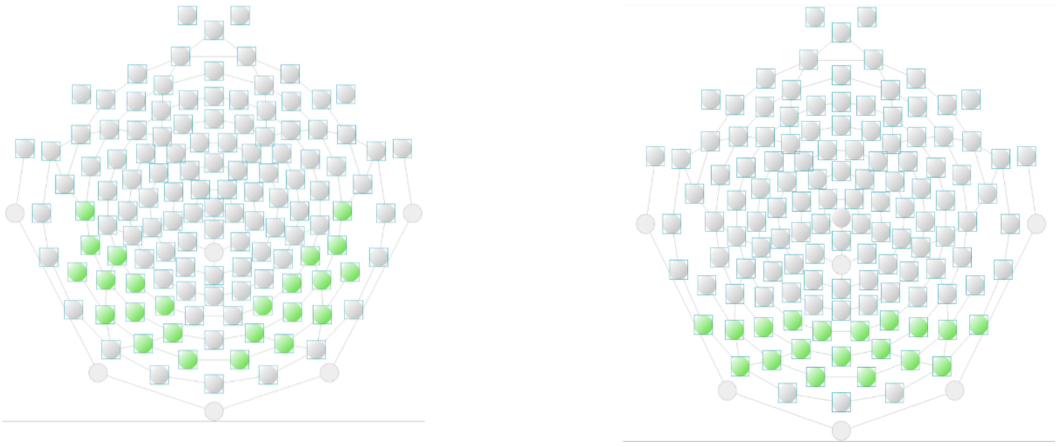

a. Dynamic gaze contrast

b. Face/noise contrast

*Figure S1.* Selected channel montages based on Elsabbagh et al. (2012) and Tye et al. (2022) corroborated with visual inspection of grand averages

### EEG Amplitude

Table SM1: *8-month-old infant ERP amplitude by mid-childhood outcome group*

| | No-autism (N=59) | | | Early-autism (N=22) | | | Later-autism (N=21) | | |
|---|---|---|---|---|---|---|---|---|---|
| | M | (SD) | n | M | (SD) | n | M | (SD) | n |
| P1 Amplitude Shift Towards | 0.61 | (2.43) | 59 | 0.41 | (2.81) | 22 | 0.31 | (2.51) | 21 |
| P1 Amplitude Shift Away | 0.97 | (2.44) | 59 | 0.14 | (3.30) | 22 | 1.07 | (1.91) | 21 |
| P1 Amplitude Towards-Away | -0.36 | (2.76) | 59 | 0.27 | (4.24) | 22 | -0.76 | (2.87) | 21 |
| N290 Amplitude to Faces | 4.40 | (6.86) | 51 | 5.69 | (6.41) | 18 | 5.83 | (6.34) | 17 |
| N290 Amplitude to Noise | 8.93 | (6.99) | 51 | 10.83 | (6.78) | 18 | 7.62 | (4.52) | 17 |
| N290 Amplitude Faces-Noise | -4.54 | (7.42) | 51 | -5.14 | (7.80) | 18 | -1.79 | (7.04) | 17 |
| P400 Amplitude Shift Towards | 1.86 | (3.75) | 59 | 2.02 | (3.25) | 22 | 1.89 | (3.84) | 21 |
| P400 Amplitude Shift Away | 2.69 | (3.20) | 59 | 1.42 | (3.98) | 22 | 3.36 | (3.29) | 21 |
| P400 Amplitude Towards-Away | -0.83 | (3.61) | 59 | 0.60 | (5.88) | 22 | -1.47 | (3.98) | 21 |

*P1 Amplitude*: There was no group, condition or group by condition interaction effect.

*N290 Amplitude:* There was a significant effect of condition ($p<.001$), but no group or interaction effect. The effect was such that the N290 amplitude was higher during the Noise versus Face condition

*P400 Amplitude:* There was no group, condition or group by condition interaction effect. The effect of condition became close to significance ($p=.051$) when removing one outlier. This effect was such that the P400 amplitude was higher to gaze shifting away versus towards.

### Associations with Autism traits

Table SM2: *Associations between ERP change scores and autism traits (Kendall Tau$_b$ (p))*

| | SRS | | ADOS | | ADI | | |
|---|---|---|---|---|---|---|---|
| | SCI | RRB | Social Affect | RRB | Social | Comm. | RRB |
| P1 Latency | .097 | .016 | .014 | .069 | .101 | .045 | .041 |
| | (.180) | (.831) | (.841) | (.370) | (.153) | (.528) | (.587) |
| P1 Amplitude | .051 | .099 | .053 | .045 | .017 | -.044 | .060 |
| | (.487) | (.185) | (.448) | (.555) | (.808) | (.532) | (.353) |
| P400 Latency | **.193** | **.214** | **.151** | **.154** | .138 | **.232** | **.169** |
| | **(.008)** | **(.004)** | **(.032)** | **(.044)** | (.050) | **(.001)** | **(.024)** |
| P400 | -.002 | -.005 | .105 | .057 | -.006 | -.027 | -.037 |
| Amplitude | (.983) | (.955) | (.133) | (.457) | (.936) | (.705) | (.622) |
| N290 Latency | -.123 | -.072 | -.041 | -.044 | -.187 | -.120 | **-.176** |
| | (.119) | (.377) | (.597) | (.604) | (.015) | (.123) | **(.031)** |
| N290 | -.035 | -.034 | .082 | .042 | .058 | .035 | .075 |
| Amplitude | (.657) | (.681) | (.287) | (.618) | (.457) | (.623) | (.360) |

Note. SRS = Social Responsiveness Scale; ADOS = Autism Diagnostic Observation Schedule; ADI = Autism Diagnostic Interview; SCI = Social Communication and Interaction; RRB = Restricted Interests and Repetitive Behaviour; Comm = Communication. Associations using Kendall Tau$_b$

### Typical Likelihood Group

We re-ran the latency models including 43 typical likelihood children. Only differences between the typical likelihood group versus other groups are mentioned here.

*P1 Latency:* There was a significant group by condition interaction effect ($\chi^2(3) = 13.47$ p=.004). The TL group differed from the early-autism group (p<.001).

*N290 Latency:* There is a significant condition ($\chi^2(1) = 7.19$ p=.007) and group by condition interaction effect ($\chi^2(3) = 9.25$ p=.026). The TL group differs from the early-autism group (p=.005).

*P400 Latency:* There was a significant group by condition interaction effect ($\chi^2(3) = 13.95$ p=.003). The TD group differed from the early-autism group (p=.006). Results remained the same after removing outlier.

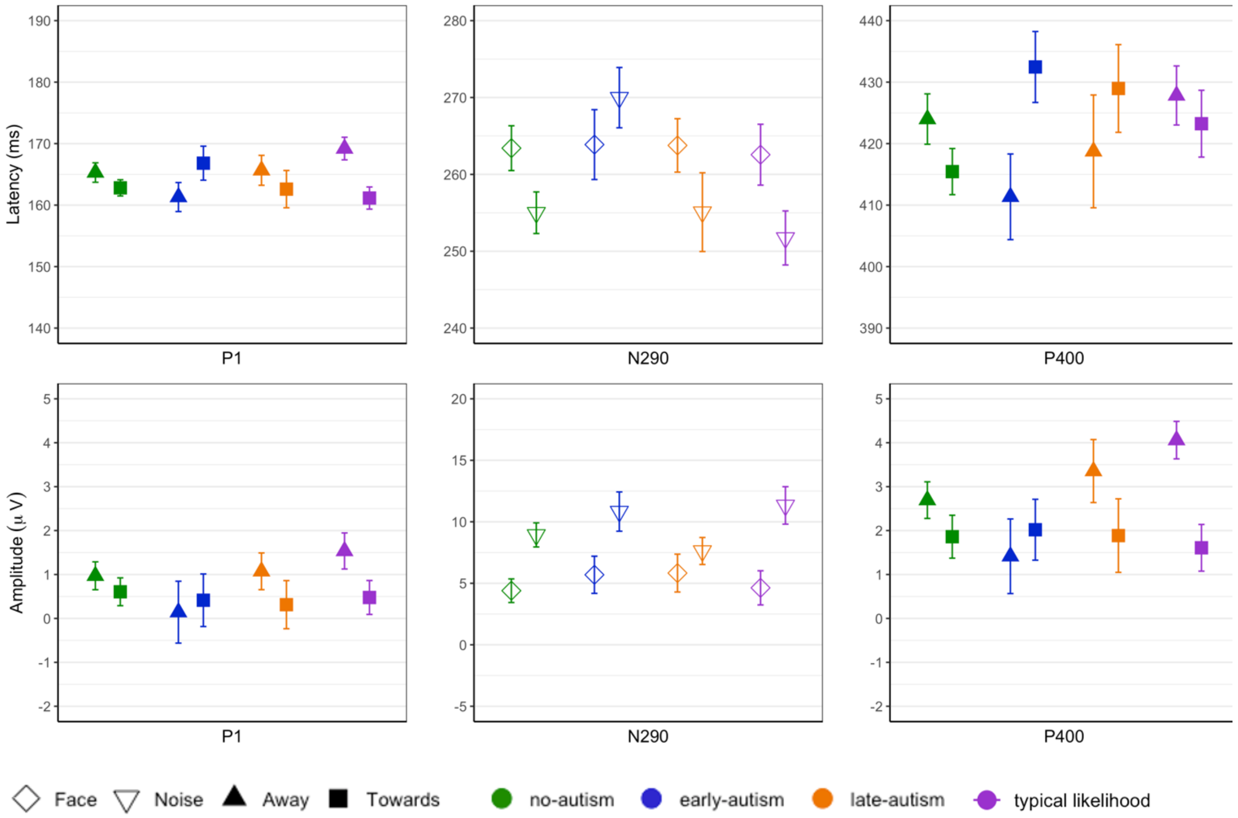

*Figure S2.* Mean and SE of latency and amplitude responses by ERP and mid-childhood outcome groups, including typical likelihood group.
